# Monitoring COVID-19 related public Interest and population Health Literacy in South Asia: An Internet Search-Interest Based Model

**DOI:** 10.1101/2020.08.24.20180943

**Authors:** Hasan Symum, Kh M. Ali Sagor

## Abstract

**Background:** Information epidemiology based on internet search data can be used to model COVID-19 pandemic progressions and monitor population health literacy. However, the applicability of internet searches to monitor COVID-19 infections and public health awareness in South Asian countries are unclear.

**Objectives:** To assess the association of public interest and health literacy in COVID-19 with the number of infected cases South Asian countries.

**Material and Methods:** Google Trends data from January to March 2020 were used to correlate public interest and literacy with official data on COVID-19 cases using the relative search volume (RSV) index. Public interest in COVID-19 was retrieved with the search topic “Coronavirus (Virus)”. Similarly, search terms “hand wash”, “face mask”, “hand sanitizer”, “face shield” and “gloves” were used to retrieve RSV indices as a surrogate of health literacy. Country-level correlation analyses were performed for a time lag between 30 and +30 days.

**Results:** There were significant positive correlations between COVID-19 related public interest and daily confirmed cases in countries expect Nepal, Bhutan, and Sri Lanka. The highest public interest in South Asian Countries was on average 12 days before the local maximum of new confirmed cases. Similarly, web searches related to personal hygiene and preventive measures in south Asia correlated to the number of confirmed cases as well as national restriction measures.

**Conclusion:** Public interest indicated by RSV indices can help to monitor the progression of an outbreak such as the current COVID-19 pandemic particularly in countries with a lack of diagnostic and surveillance capacity.

## Introduction

The coronavirus disease 2019 (COVID-19) pandemic represents an unprecedented global healthcare emergency with more than 20 million laboratory-confirmed cases and 730,000 deaths between February and July 2020 [1]. COVID-19 is caused by the novel acute respiratory syndrome coronavirus 2 (SARS-CoV-2 and first reported in Wuhan, China on December 31, 2019 [2]. Since December 2019, the outbreak continues to spread rapidly worldwide due to the high viral transmissibility and prevalence of asymptomatic cases [3,4]. By March 2020, COVID-19 has spread globally with more than 400,000 confirmed cases which prompted the World Health Organization (WHO) to declare the COVID-19 as a global pandemic [5]. As of August 10, 2020, COVID-19 has affected more than 227 countries and territories with more than 7 million active cases and the number is still exponentially rising [1,6]. South Asia is one of the leading COVID-19 affected regions with more than 20 million (28.57 % of global cases) confirmed cases as of August 2020. India, Pakistan, and Bangladesh were the most COVID-19 affected countries in the South Asia region with 1695988, 278305, and 237661 confirmed cases, respectively [6].

Over the last few years, the internet has become the principal source of information particularly for any healthcare-related concerns. [7,8]. With over 4.5 billion active internet users around the world, millions of people worldwide search online for health-related queries which make Web search queries a valuable source of public health infoveillance [9,10]. Understanding about Web search trends can provide valuable information about public interest and awareness in health emergencies as a proxy for public health risk perception [8,11]. Prior studies used internet search queries to model the outbreak of infectious disease (e.g., dengue, and influenza), track substance usages, and monitor public behavior [12,13]. Internet search queries also used to investigate public interest, health awareness, and mental health in the COVID-19 pandemic situation [14–16]. Most of these studies focused on China, the Americans, and other European countries. However, infodemiology studies exploring the association of web queries to the COVID-19 public interest and awareness in South Asia received limited attention. So far, only one study showed promise in exploring the potential use of Google Trends in predicting the COVID-19 outbreak in India, and yet, the relationship between web queries and public health preparedness remained unclear [17]. In addition, there is no Infodemiology study till now that has explored the association of web queries to the COVID-19 public internet and awareness in other seven South Asian countries. With more than 880 million (18.87% of global users) internet users in South Asia, tracking Web search queries can be a real-time health informatics tool to strengthen the public health surveillance in health emergencies such as COVID-19 pandemic [9,10,18]. Therefore, this study explored the potential use of internet search trends for monitoring public interest and preventive health awareness towards COVID-19 infections in South Asia.

## Methods

Daily data on confirmed COVID-19 cases were obtained from the data repository managed by the Center for Systems Science and Engineering (CSSE) at Johns Hopkins University [6]. Data were retrieved for worldwide total new cases as well as for the following individual countries namely the United States (US), Afghanistan, Bangladesh, Bhutan, India, Maldives, Nepal, Pakistan, and Sri Lanka from the 24th January 2020 to 29th July 2020.

The Google Trends analytics platform was utilized to explore internet user search activities related to COVID-19 pandemic public interest and population health literacy. Google Trends is an online tracking system of internet hit-search volumes, which enables the researchers to study trends and temporal patterns of the popular search queries [19]. Google trend determines the proportion of the searches for user-specified terms among all web queries on the Google Search website and other affiliated Google sites for a given location and time [20]. Google trend then normalized the proportion by the highest query share of that term over the time-series and reports search interest as a unit of relative search volume (RSV) index. The retrieved RSV index values range from 0 to 100, with a value of 50 representing half the public interest as a value of 100 [21]. This RSV indices have been used previously used to analyze public interest in various health-related issues as well as passive health surveillance and disease monitoring [13,14,22]. The search topic “Coronavirus (Virus)” was used to retrieve Worldwide and nation-specific RSV indices in Google Trend as a representation of public interest on COVID-19 information. Also, a OR combination of the popular search phrases “hand wash”, “face mask”, “hand sanitizer”, “face shield” and “gloves” were used to retrieve RSV indices as a surrogate of public interest on the practice of personal hygiene and other COVID-19 preventive measures.

Changes in temporal trend of public interest and number of COVID-19 cases were analyzed graphically for nation-specific major events and policy initiatives (e.g., first coronavirus cases,1000 cumulative deaths, nationwide lockdown). Temporal association in public interest in COVID-19 and the number of new confirmed cases were analyzed using time-lag correlations measures. The lag correlation assesses whether the increases in GT data were correlated with the subsequent increase in COVID-19 cases and how far (days) the two series are offsets [23]. Pearson correlation coefficient was used to correlate between public interest and COVID-19 cases for a time lag between 30 and +30 days similar to prior studies [22,24–26]. We used Pearson correlation based on our assumption of linear relationship between the public interest and COVID-19 cases. Data of south Asian countries were also compared with the United States which was one of the most affected countries by the COVID-19 pandemic to investigate whether these searching terms objective for other communities or not. For the web interest and confirmed new case data, the period was set from January 24 to July 30, 2020 for the worldwide and induvial countries. For the countries’ time lag and correlation analysis, COVID-19 cases, and RSV indices data from January 24 to April 30 were used. Initial cutoff date of January 24 was selected as the day after first COVID-19 confirmed case in South Asia on January 23, 2020 [27]. April 30 was selected as the ending cut off point approximately one month after the lockdown implementation and rapid disseminations of COVID-19 related information. Each country’s data was examined individually, and no direct comparison was made between countries in COVID-19 data or RSV index data. For each time frame, a new google trend dataset for was retrieved and matched with the official COVID-19 confirmed case data for further analysis. All the statistical analyses were performed using R studio and p value less than 0.05 was considered significant. IRB approval was not required because this study did not involve human subjects.

## Results

Worldwide public interest in coronavirus started to increase from January 22nd,2020 and reached its first local peak on January 31, 2020, when WHO declared Covid-19 as a national healthcare emergency (Figure 1). Then COVID-19-related worldwide searches remain low for some time and continuously increased after the word was spread on the outbreak in Wuhan, China. Worldwide COVID-19 related searches. COVID-19-related worldwide public interest continued to expand and reached a peak on March 16, 2020, as worldwide coronavirus cases and related death were reported and right after the WHO announcement to declare the coronavirus outbreak as a pandemic. Worldwide COVID-19 related searches remained steadily high for 1 week and again reached a peak on March 22, 2020, after the massive spread of coronavirus around the world, particularly in European and American countries. There are two small peaks, one sharp increase in numbers on February 12, 2020, when china adjusted coronavirus cases from confirmatory laboratory test and the other peak on April 12, 2020, due to cases around the globe. As of 29 July 2020, the daily number of confirmed coronavirus cases still increasing almost every day and has not reached its highest point yet. The number of reported confirmed COVID-19 cases in Bhutan, Afghanistan, Maldives, Nepal, and Sri Lanka were lower compared to other South Asian countries. India, Pakistan, and Bangladesh were the most COVID-19 affected three South Asian countries that overtook China in terms of the number of coronavirus cases. The COVID-19 related searches in South Asia reached its initial peak right after the 1st confirmed case reported in the most countries (Figure 2). After the first peak, public interest in COVID-19 related information increased rapidly after the dissemination of confirmed cases in mainland China and Europe. Public searches about the coronavirus in these countries reached its second major peak right after the announcement of strong lockdown measures and a few days after the numbers of newly locally confirmed COVID-19 started to increase exponentially. COVID-19 related searches reached the highest peak approximately a few days after each country reported 100th confirmed cases except for Bhutan and Maldives. Then public interest in South Asian countries remained almost steady the same for two-three weeks with rapid increases in the number of COVID-19 cases. After the end of late April, COVID-19 related searched continuously declined similar to worldwide trends. The daily number of confirmed COVID-19 cases in most countries reached a peak in late Jun and early July 2020 except for the Maldives, where daily COVID-19 cases peaked on 30th April 2020. The daily number of confirmed COVID-19 cases has dipped from peak level in any South Asian Counties except India (Table 1).

**Table 1:** Timeline Of COVID-19 Progression and National Restriction Measures.

| Country | Confirmed 1st case (date) | Total 1000 cases (date) | Total 10,000 cases (Date) | Highest daily cases | lockdown measures (date) | Lockdown eases (date) | Total Confirmed cases (07-30-20) |
| --- | --- | --- | --- | --- | --- | --- | --- |
| Afghanistan | 24 Feb-20 | 20 Apr-20 | 24 May-20 | 05 June-20 | 14 Mar-20 | 21 May-20 | 36675 |
| Bangladesh | 08 Mar-20 | 14 Apr-20 | 04 May-20 | 02 July-20 | 23 Mar-20 | 30 May-20 | 237661 |
| Bhutan | 20 Mar-20 | - | - | 06-June-20 | 22-Mar-20 | - | 101 |
| India | 30 Jan-20 | 29 Mar-20 | 14 Apr-20 | - | 25 Mar-20 | 31 May-20 | 1695988 |
| Maldives | 08 Mar-20 | 15 May-20 | - | 30 Apr-20 | - | - | 3793 |
| Nepal | 23 Jan-20 | 28 May-20 | 23 June-20 | 07 July-20 | 24 Mar-20 | 24 July-20 | 19771 |
| Pakistan | 26 Feb-20 | 24 Mar-20 | 21 Apr-20 | 06 June-20 | 24 Mar-20 | 09 May-20 | 278305 |
| Sri Lanka | 27 Jan-20 | 19 May-20 | - | 10 July-20 | 20 Mar-20 | 11 May-20 | 2815 |

**Figure 1:**
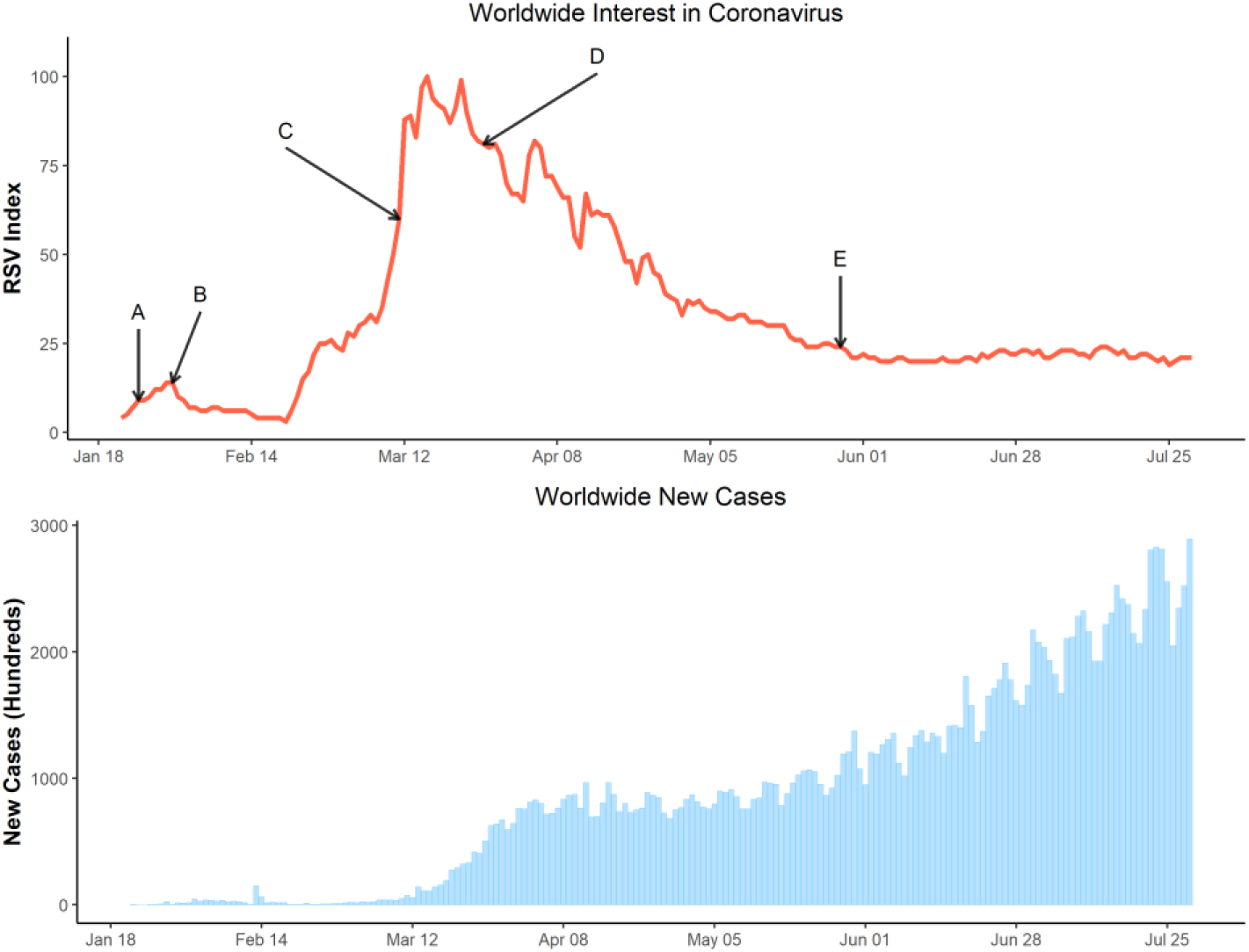
Time series of relative search volumes (RSVs) related to COVID-19 and worldwide daily cases. Note, A: 1000 case reported worldwide [January 25,2020], B: WHO declared health emergency [January 31,2020], C: WHO declared COVID-19 as pandemic [March 11,2020], D: 1000 death reported in USA [March 26, 2020], E: 100000 death reported in USA [May 28, 2020]

**Figure 2:**
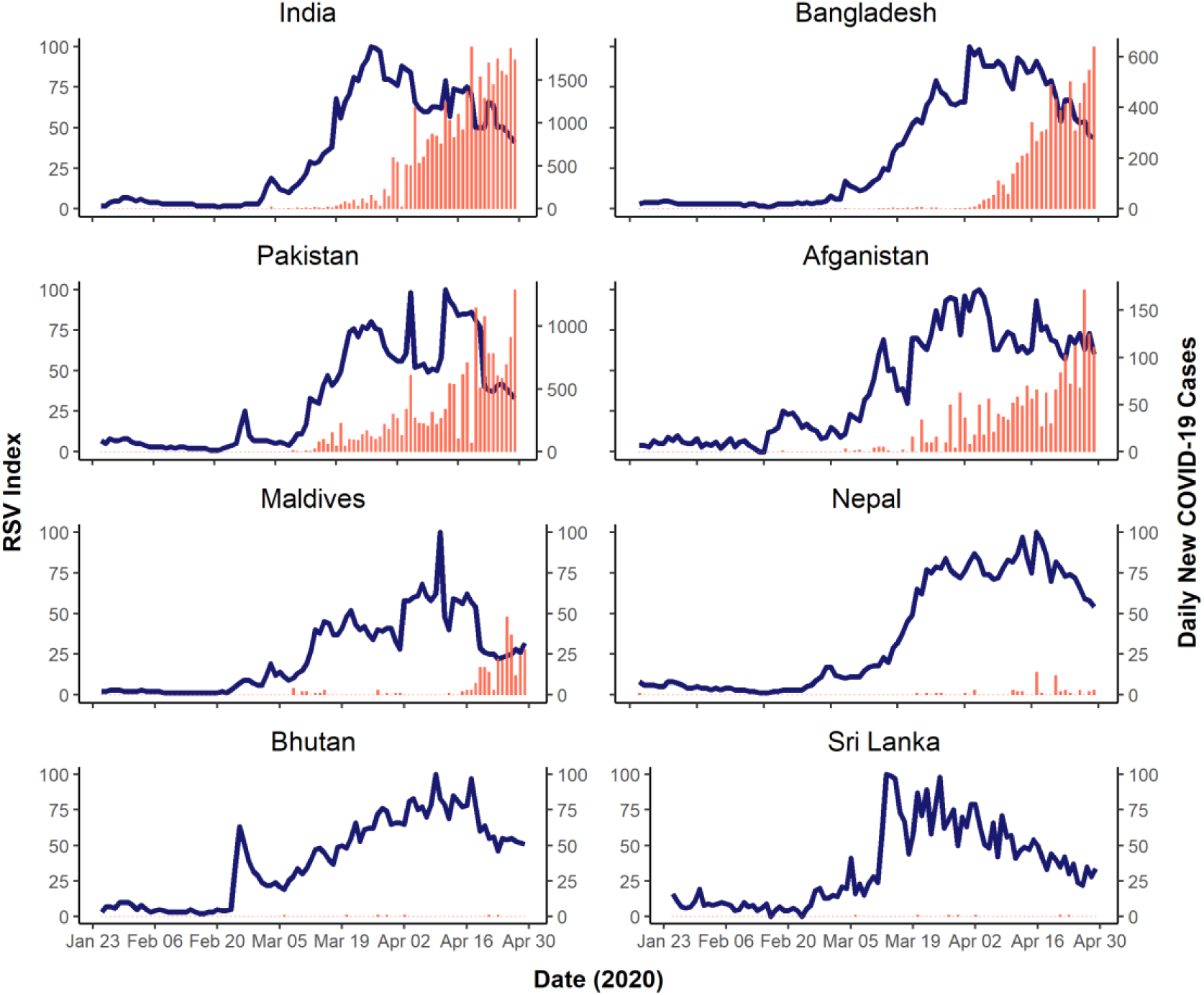
Time series of relative search volumes (RSVs) related to COVID-19 and daily cases in South Asian countries

From the correlation analysis, the highest public interest in COVID-19 was on average 9 days before the local maximum of new confirmed cases (Figure 3). Our study found a significant high correlation (> 0.70) in Bangladesh, Pakistan, India, and Afghanistan, where the Maldives showed a moderate (between 0.50 and 0.70) time lag correlation. A weak time-lag correlation (< 0.5) was found between searching patterns and disease outbreaks in Bhutan, Nepal, and Sri Lanka. India and Bangladesh show a similar pattern with the highest correlation between RSV indices and newly confirmed cases found with time lags of −16 and −12 days, respectively. In addition, Pakistan shows a similar positive correlation pattern with the highest coefficient found with a slightly shorter time lag of −9 days than the two countries. All three countries show comparable patterns in correlation coefficient with the US, where the highest coefficient found with a slightly longer time lag of −19 days. According to Figure 3, the dynamics of web searches in South Asia related to personal hygiene and COVID-19 preventive measures were related to vulnerability due to confirmation of local transmission of COVID-19 and public restlessness given the announcement of national restriction measures.

**Figure 3.**
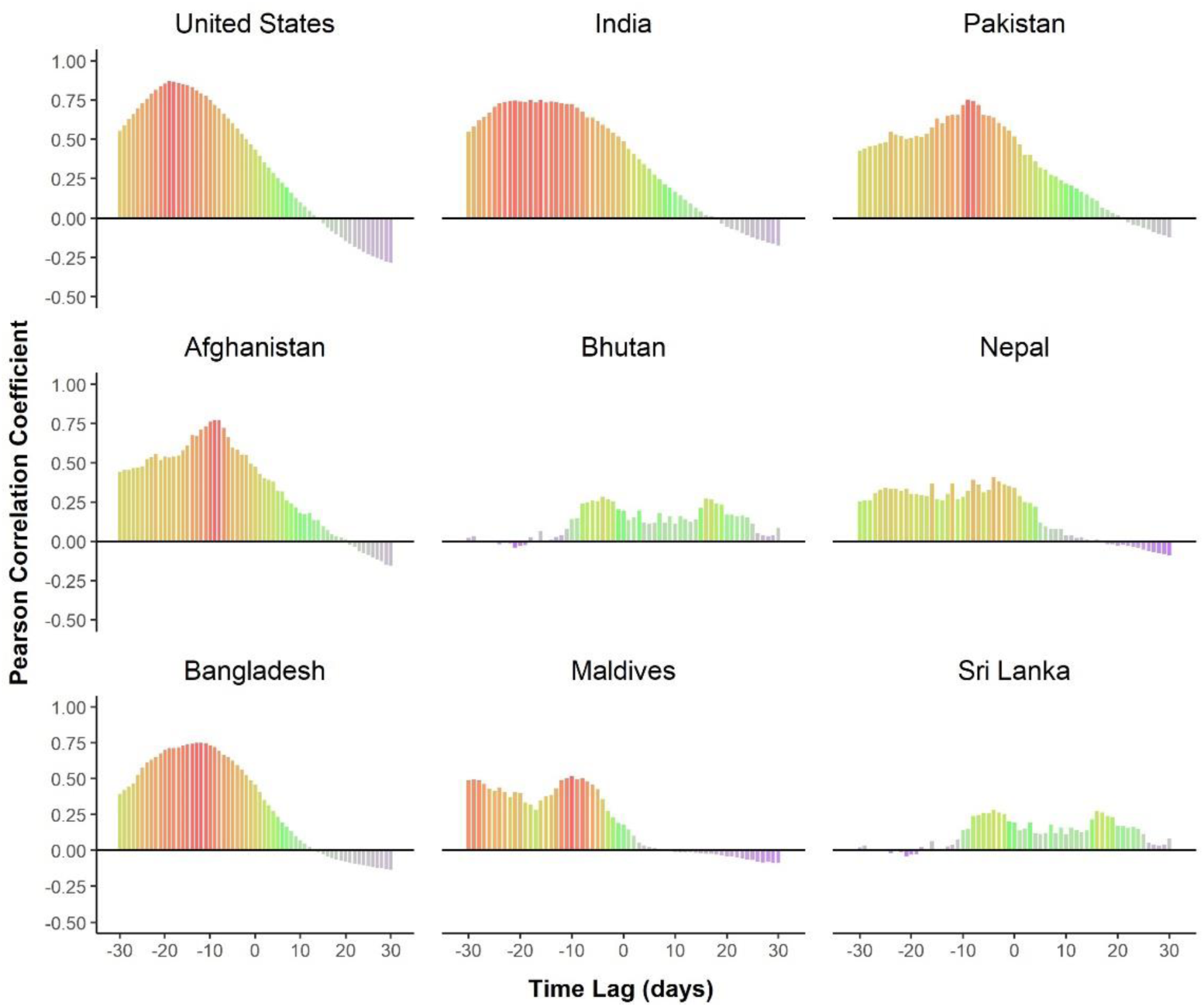
Time-lag correlations of Relative Search Volume (RSV) indices for COVID-19 and daily new confirmed COVID-19 cases for South Asian Countries

**Figure 4:**
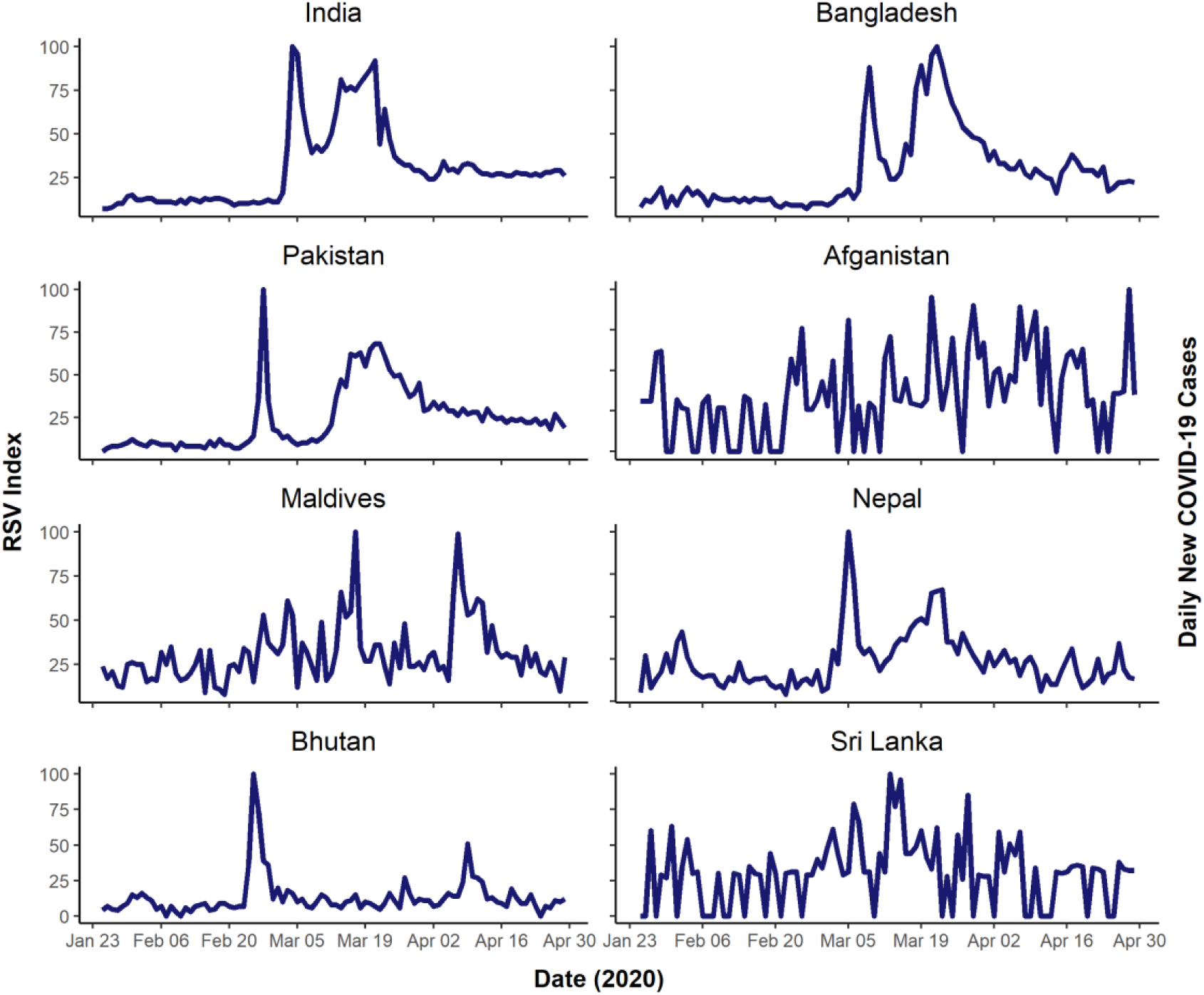
Time series of relative search volumes (RSVs) related to COVID-19 health literacy and daily cases in South Asian countries

## Discussion

Over the last few months, the coronavirus pandemic has become a global public health crisis and an unprecedented disruption of daily life around the world. People around the world, particularly in the affected countries, have faced extreme uncertainty due to evolving knowledge regarding COVID-19 symptoms and corresponding novel behavioral changes to prevent community transmission. In this study, we found substantial changes in coronavirus information-seeking countries all over the world, particularly in March 2020, suggesting a hyper-awareness and mass interest for information about COVID-19 pandemic. In addition, we found a varied level positive correlation between public interest and real-world cases in South Asian countries, with internet searches preceding real-world cases by on average 9 days. This lag time reflects the increases in the public attention weeks before the increases in COVID-19 cases occurred in most South Asian counties. This lag time biases further justices the importance of pursuing more real-time assessments of COVID-19 pandemic growth when there is an increase in internet activities as people develop symptoms [16,25]. The longer the maximum time lag represents the earlier search engines were able to predict the COVID-19 outbreak. The maximum time lag period in India for COVID-19 cases and public interest was found to be −16 days that is, the search volume for “coronavirus” peaked 16 days before the peak number of cases. The longer lag time for the US compared to countries in South Asia may be attributed to the sensational dissemination of COVID-19 news from China and other European countries, which could have influenced their web search behavior. These findings indicate that internet query searches can be a useful tool to predict and monitor the outbreak of infectious diseases, thus can provide a valuable picture of the outbreak of COVID-19 pandemic progression in real-time.

Close monitoring and continued evolution of enhanced communication strategies are urgently needed to provide general populations and vulnerable populations with actionable information for self-protection and clear guidance during an outbreak [28]. The application of electronic medium, specifically the internet data in health care research, known as infodemiology, is a promising new field that provides unmatched opportunities for the management of health information generated by the end-users [10]. Using this unique potential, previous researchers were able to correlate the internet searches with traditional surveillance data and can even predict the outbreak of infectious disease several days or weeks earlier [11,13]. Recent COVID-19 related infodemiology studies modeled daily laboratory-confirmed / suspected cases and associated death with internet search queries in the US, China, Iran and several European countries [16,18–20,25,29]. Also, the regression/ machine learning model using the Google search queries moderately predicted the incidence of COVID-19 in Iran. Similar attempts were subsequently made to predict the previous coronavirus related outbreaks (e.g., SARS, MERS) and other infectious diseases (e.g., Zika, dengue) [11,13,30]. The fundamental assumption behind this disease monitoring using google trends is that mostly symptomatic and soon to be symptomatic people and people will search about the disease on the internet before accessing healthcare [31]. In our study, we assumed the majority of the COVID-19 infections are symptomatic, and people will search for information regarding COVID-19 and associated preventive measures. A recent study estimated that the majority (80%) of people with an infection with the new coronavirus showed any types of symptoms while others (20%) remain symptom-free [32].

Despite the studies above, there is still a limited of research COVID-19 infodemiology study in South Asia. According to WHO, South Asia is considered highly vulnerable to any large-scale outbreak of an infectious disease being one of the most populous word region [33]. Therefore, the use of infodemiology particularly internet searches provides unique opportunities to monitor real-time public interest and awareness, particularly in countries with a lack of diagnostic and surveillance capacity, and thereby disseminate evidence-based health information to the people [34]. In South Asia, the first imported coronavirus case was reported in Nepal on 23 January 2020 [6]. Starting from the March 2020 number of coronavirus cases started to increase rapidly in South Asian countries except for Bhutan (Table 1). Countries in the region instituted various levels of national restriction measures and ban on international travel starting from the mid-March 2020 [15]. The number of reported confirmed COVID-19 cases in Bhutan, Maldives, Nepal, and Sri Lanka were at a low compared to other South Asian countries. These countries are either completely sea locked or sparsely populated and completely sealed their international border, which may have contributed low COVID-19 infections compared with their neighbors [35,36]. Besides, weak correlation (< 0.5) between searching patterns and disease outbreaks in Bhutan, Nepal, and Sri Lanka might be related to the lower number daily of COVID-19 cases than other countries in South Asia from January 24 to April 30. According to the WHO’s report on the 11th March 2020, that the outbreak of COVID-19 meets the criteria for a global pandemic and people’s interest started to increase in all south-Asian Countries. However, People in India and Pakistan had a faster response speed toward COVID-19 than Bangladesh which is likely to be related to the number of local cases and lower virulence to COVID-19. As of the 15th March 2020, the number of patients confirmed with COVID-19 in Bangladesh (05 cases) was lower than in India (110 cases), and Pakistan (31 cases) [6]. After the peaks within two-three weeks, internet searches continued to decline declined due to massive dissemination of information reported on the local/national news reporting, video news reporting, and health expert reporting in social media [16,18]. These findings suggest that internet searches can potentially help governments to define proper timing of risk communication, improve the public’s vigilance, and strengthen the publicity of precautionary measures when facing any public health emergencies like COVID-19 [37].

This study has some limitations that should be acknowledged. First, this study used single search engines, Google, to retrieve population interest data form the South Asian Countries. Thus, there might be selection bias since people who use other search engines are not included in this investigation. However, since more than 880 million people in South Asia use the internet and google as the major search engine (more than 98% market share), google search queries can be a strong tool to estimate public interest [9,38]. Second, Google trend do not report search query result the form of a relative search value instead of absolute search volumes which might have limited more in-depth and precise investigations. In addition, the Google trend excludes all the search results with any typographical error in the query terms. Third, although the number of studies based on google trend is increasing, Google does not provide the detailed information about the procedures by which they generate search data, and the study population responsible for the searches remain unclear [13]. Finally, search volumes can be influenced by the dissemination of information through the news media, and it is still unclear whether changes in online activity translate to changes in health behavior. This massive dissemination might have influenced RSV values through disproportionate swings among the citizens in different countries and may have resulted in overestimation about public health literacy and public awareness. This requires caution when analyzing results and making interpretations from the analyses. Therefore, it is strongly recommended that Google trends should not be used as a sole replacement for robust disease surveillance and health literacy monitoring, rather it should be a real-time supplement to the comprehensive disease monitoring [17].

## Conclusion

COVID-19 related worldwide public interest reached a peak on March 16, 2020, right after the WHO announcement of COVID-19 as a pandemic. COVID-19 related public interest reached the peak in South Asian countries within a few days after reporting 100th confirmed cases in each country. Significant positive correlations between COVID-19 related public interest and daily laboratory-confirmed cases were found in most Countries. The average time lag of 9 days suggests the increment in the public attention weeks before the increases in COVID-19 cases occurred in most South Asian counties. Therefore, infodemiology studies particularly using internet search volume provides an unorthodox opportunity to monitor the real-time progression of an outbreak such as COVID-19 and associated population health interest in countries particularly with limited surveillance capacity, and thereby distribute health information to the public.

## Data Availability

Data is publicly available at https://trends.google.com/trends/?geo=US

